# City-wide Built Environment SARS-CoV-2 Detection for COVID-19 Surveillance

**DOI:** 10.1101/2025.03.06.25323509

**Authors:** Derek R. MacFadden, Michael Fralick, Caroline Nott, Jason A. Moggridge, Alexandra M.A. Hicks, Tamara Van Bakel, Evgueni Doukhanine, Aaron Hinz, Nisha Thampi, Pascal Bastien, Sarah Mansour, Engluy Khov, Tasha Burhunduli, Douglas G. Manuel, Alex Wong, Rees Kassen

## Abstract

Built environment surveillance has shown promise for monitoring COVID-19 burden at granular geographic scales, but its utility for surveillance across larger areas and populations is unknown. Our study aims to evaluate the role of built environment detection of SARS-CoV-2 for the surveillance of COVID-19 across broad geographies and populations. We conducted a prospective city-wide sampling study to examine the relationship between SARS-CoV-2 on floors and COVID-19 burden. We used non-parametric correlation analyses and linear models to evaluate associations between SARS-CoV-2 signals and COVID-19 outcomes, across multiple locations/populations over time. Sampling sites included schools, libraries, and emergency departments in the capital city of Ottawa, Canada, from October 2022 to March 2023. Floor sampling was performed across spaces, and outcomes were evaluated at aggregate levels. Detection (presence/absence) and quantification (viral load) of SARS-CoV-2 was determined by reverse-transcriptase polymerase chain reaction conducted on floor swabs collected weekly at study locations. The main outcomes, and measures of COVID-19 burden, were (1) weekly regional wastewater signal and (2) weekly admitted COVID-19 patient census from hospitals. We collected 1,863 built environment floor samples over the 6-month study period, with an overall swab positivity for SARS-CoV-2 of 45% (95%CI 43%-48%). We found a strong correlation between overall built environmental swab viral load and hospital COVID-19 census (Spearman’s *r*=0.64, *p*=0.0017), but no correlation between regional wastewater and hospital COVID-19 census (Spearman’s *r*=-0.15, p=0.5). We found a strong correlation (Spearman’s *r*=0.76, *p*=9x10^-5^) between hospital-specific swab viral copy number and hospital-specific COVID-19 census, which is a likely driver of the overall association between swab load and census. Built environment surveillance of SARS-CoV-2 from hospitals was strongly correlated with hospital burden with improved delineation of hospital COVID-19 cases compared to regional wastewater. These findings support the use of built environment surveillance for quantification of infectious burden amongst institutionalized groups.

## INTRODUCTION

The COVID-19 pandemic has had major impacts on human health globally.^1^ Over the five years since the first identification of SARS-CoV-2 (the agent of COVID-19), environmental surveillance in the form of wastewater testing has emerged as a crucial public health tool for monitoring population burden (particularly when testing cannot be captured accurately by health systems, e.g., from rapid antigen tests) and for anticipating changes in population burden.^2^ The World Health Organization and the United Nations have recognized the importance of environmental surveillance for SARS-CoV-2, and highlighted the necessity of its role moving forward for measuring and anticipating the burden of COVID-19 and other pathogens of concern.^3,4^

While wastewater has been the dominant form of environmental SARS-CoV-2 surveillance, built environment detection has also emerged as a complementary tool, with appealing characteristics that include: (1) low capital cost; (2) ease of use; (3) lack of requirement for wastewater infrastructure; (4) flexibility to detect non-GI excreted pathogens; and (5) high spatial resolution.^5–7^ Built environment detection is a type of environmental surveillance which is focused on sampling from built (human constructed) surfaces which exist in both urban and rural environments. Built environment detection has been evaluated for a growing number of SARS-CoV-2 surveillance indications, from hospitals to nursing homes to schools.^5–8^ In a large prospective multi-center observational study of 10 nursing homes, built environment sampling of SARS-CoV-2 was able to anticipate COVID-19 outbreaks, sometimes by 1-2 weeks.^5^ Built environment detection of SARS-CoV-2 also parallelled case burden and wastewater signal at a large tertiary care hospital.^5,7,9^ Most evaluations to date of built environment surveillance for SARS-CoV-2 have been focused on spatially refined locations and populations.^5,7,9^ Here, we propose that distributed built environment sampling of locations/populations over a large region may facilitate broader surveillance, which could augment or potentially supplant other forms of surveillance. Such an approach could be particularly useful for areas without existing wastewater surveillance (e.g., remote communities), or to monitor COVID-19 burden in high-risk populations (e.g., institutionalized patients), as subgroup groups of broad surveillance efforts. ^5,7,9^

The objective of our study was to evaluate the use of built environment screening for SARS-CoV-2 as a measure of COVID-19 burden across a large metropolitan city. In particular, we sought to determine whether SARS-CoV-2 detection could provide relevant population and neighborhood-specific burden estimation.

## MATERIALS AND METHODS

### Study Design

We performed a prospective city-wide built environment sampling study across six geographically distinct regions in the city of Ottawa, Canada (Supplemental Figure 1) to evaluate the relationship between environmental sample results and relevant COVID-19-related outcomes, including hospital case burden and wastewater SARS-CoV-2 signal. Sampling regions were selected *a priori* based upon their unique populations, vulnerable groups, and sewersheds. For these regions, we selected separate neighbourhoods that contained the key populations identified below, and also the relevant corresponding sampling environments where possible. Sampling environments were chosen to represent key populations, including school sampling (school-aged children), public library sampling (healthy adult residents), and ED waiting and assessment areas (ill or vulnerable individuals). Floor sampling was performed once weekly at each site from every geographic location, from October 2022 to March 2023.

### Ethics

This study was performed under an REB exclusion from the University of Ottawa, given that it did not involve individuals or individual-level data.

### Sample Collection

We received permission from representatives at each built environment site to collect our samples. We performed sampling at five large tertiary/quaternary care hospitals, six Ottawa Public Library (OPL) branches, and five publicly-funded elementary schools from the City of Ottawa. Prior to the initiation of swabbing, we conducted site visits to select floor locations to sample, and these were selected with the intent of capturing areas with moderate to high traffic (e.g., ED waiting rooms, common meeting areas of schools, and shared spaces in libraries). A single swab was collected per location, typically between the hours of 8 a.m. and 6 p.m. Hospitals and libraries had five to six floor swabs collected per week, and schools had eight to 10 floor swabs collected per week. Swabbing was performed weekly over the study period in the same approximate areas based on previously established methods.^9^ In brief, the sample collection process went as follows: swabs were (1) wet with nucleic acid stabilization solution; (2) passed repeatedly across an approximate 2” x 2” area for 30 to 60 seconds; and (3) stored in a nucleic acid stabilization buffer for transfer to the central laboratory. Swab kits consisted of the P-208 Environmental Surface Collection Prototype flocked swabs and swab storage vials, provided in-kind by DNA Genotek.

### Sample Processing and Analysis

Swab intake, nucleic acid extraction, and reverse-transcriptase polymerase chain reaction (RT-qPCR) were performed at a local central laboratory to identify the presence of SARS-CoV-2, as previously described.^9^ Both binary detection (positive/negative) and quantitation cycle (Cq) were determined for each swab. A Cq cutoff of 45 was used to determine whether a swab was positive or negative^9^. Cq was converted to viral copies (viral copies + 1 to facilitate log scale) using previously-determined standard curves.^9^

### Exposure Variables

Exposure variables included (1) percent positivity of built environment swabs for a given week and (2) geometric mean viral copies plus one (using standard-curve based relative quantification from cycle thresholds) of built environment swabs for a given week. One copy is added to each value, such that non-detection of SARS-CoV-2 is represented as one copy to ensure that all values are log-transformable and included in aggregate statistics for viral copy numbers.

### Outcome Variables

We evaluated the following outcomes: (1) weekly average level of combined SARS-CoV-2 N1 and N2 targets from City of Ottawa wastewater collected at the Robert O. Pickard Environmental Centre (relative to pepper mild mottle virus - PMMoV),^10^ and (2) seven-day midpoint median number of COVID-19 patient census from participating hospitals. A seven-day midpoint median was used to provide a measure of central tendency that could be used to compare with weekly swabbing results, During the time of our study, COVID-19 testing by PCR was not recommended except for select populations at risk of severe disease.^11^ As a result, COVID-19 case counts were not available for the non-hospitalized populations under study (i.e., schools and libraries). The results of regional wastewater testing at the Robert O. Pickard Environmental Centre in Ottawa were acquired from the Zenodo repository.^10,12^ Seven-day midpoint moving averages were computed from the daily wastewater testing results for correlation analyses with the results of surface swabbing.

### Neighborhood Grouping

Collection sites were grouped into neighborhoods, with each neighborhood having one hospital, one school, and one library under surveillance (with the exception of Neighborhood-2, which included solely a library under surveillance).

### Statistical Analysis

Results of swab tests were grouped into weekly bins spanning from Sunday to Saturday, with the midpoint occurring on the Wednesday, to compute the weekly mean swab positivity rate and the geometric mean of viral copies, and 95% confidence intervals on these means. Confidence intervals on sample proportions were calculated by Wilson’s method. Confidence intervals on sample means were calculated using the student-t distribution. Weekly aggregate statistics were computed at each of the sites, facility-types, and neighborhood levels (i.e., multiple sites in various settings located in proximity to each other). Weekly SARS-CoV-2 detection rates and quantification were compared with covariates (wastewater signals and hospital census) over time through graphical analyses and in two-sided Spearman’s (and Pearson’s where indicated) correlation tests.

We modeled the per-hospital weekly average number of patients currently admitted with COVID-19 (census) using a linear model (time-invariant) with random intercepts for each hospital building, accounting for the repeated measures on these observational units. We examined three different models: one with the weekly swab test positivity as a fixed effect, one with the per-swab viral load (i.e., weekly mean log-10 transformed copies) as a fixed effect, and one with both these measures as fixed effects. The general formula for these mixed effect models is as follows:

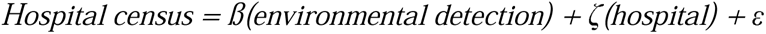

Where ß represents the coefficients for environmental detection variables modeled as fixed effects, ζ are random intercepts for each hospital (to account for repeated measures on these observational units), and ε is random error. Mixed effects models were fitted using the “lmer” function provided by the “lme4” package for R. Each model specification was evaluated through 5-fold cross validation (repeated 10 times), with the resampling procedure being stratified by site and performance estimated in terms of root-mean-squared error (RMSE) and the coefficient of determination (R^2^). All analyses were performed in the R programming language (v4.3.2).^13^ STROBE guidelines were followed for reporting observational studies where applicable.^14^

As a sensitivity analysis to evaluate whether there may be delayed responses between built environment detection and burden measures, we also explored lagged relationships between environmental detection and hospital census.

### Role of Funders

This project was supported by funding from a CoVaRR-Net Rapid Response Research Grant (#ARR-175622). The funders had no role in the design, data collection, data analysis, interpretation, or writing of the report.

## RESULTS

We collected 1,863 swabs from 16 locations in Ottawa, Ontario, Canada, between October 25, 2022 and March 30, 2023. Overall, 846 swabs were positive for SARS-CoV-2 (45%, 95% Confidence Interval [CI]: 43-48%) and the geometric mean number of viral copies per swab was 3.35 (95% CI: 3.12-3.61).

### SARS-CoV-2 Detection By Setting/Population

We examined the variability of SARS-CoV-2 built environment detection from different congregate settings, including hospitals, libraries, and schools. We found that there were large differences in SARS-CoV-2 detection rates between settings (Table 1), with hospitals having the greatest positivity at 92% (95% CI: 89-94%), while libraries (31%, 95% CI: 28-35%) and schools (29%, 95% CI: 26-32%) had more moderate detection rates. Average viral quantification followed a similar trend, with swabs collected at hospitals capturing a greater quantity of viral RNA (geometric mean 23.7 copies, 95% CI: 20.5-27.3), compared with schools (1.71 copies, 95% CI: 1.6-1.83) and libraries (1.82 copies, 95% CI: 1.69-1.96).

**Table 1.**
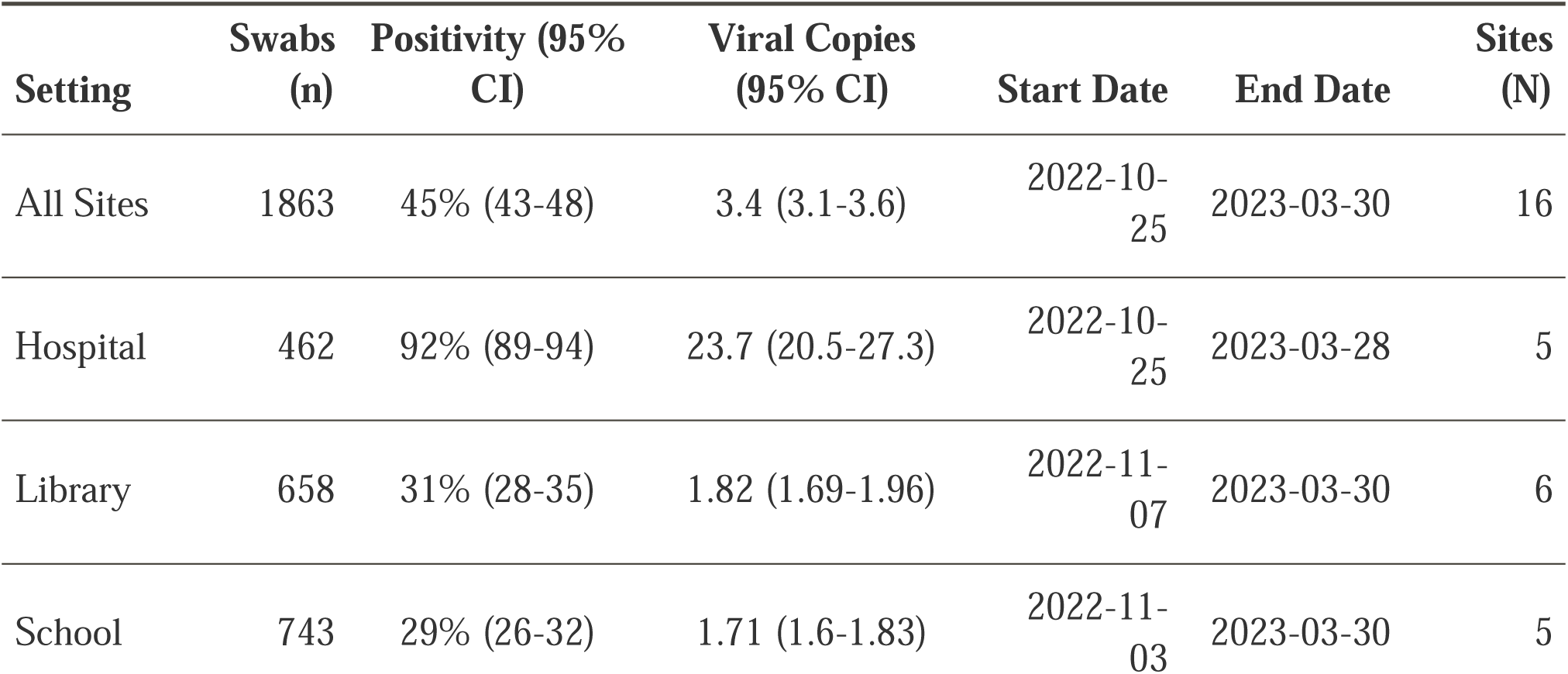
Descriptive statistics by setting/population.

### SARS-CoV-2 Detection by Neighborhood

We sought to describe the geographic variability of SARS-CoV-2 built environment detection across 6 neighborhoods, with collection sites following our grouping strategy (see Methods). Collection sites were grouped together based on proximity, with each neighborhood being associated with approximately one hospital, one school, and one library under surveillance (with the exception of Neighborhood-2, which had solely a single library under surveillance). All neighborhoods with multiple sites under surveillance had similar rates of swab positivity (Supplemental Table 1), with the greatest positivity occurring in Neighborhood-4 (54%, 95% CI: 48-59%), and the lowest in Neighborhood-5 (41%, 95% CI: 36-46%). In terms of viral RNA quantities (geometric mean), Neighborhood-4 again had the greatest amount among neighborhoods (4.96 copies, 95% CI: 4.11-5.99); Neighborhood-3 also had a similarly large viral load (4.72 copies, 95% CI: 3.84-5.8). Swabs collected in Neighborhoods-1, 5, and 6 had significantly lower quantities of viral RNA than Neighborhoods-4 and 3.

### SARS-CoV-2 Detection Over Time

We evaluated trends in SARS-CoV-2 swab positivity over time across hospitals, libraries, and schools (Figure 1). Libraries and schools showed a similar pattern, both demonstrating a peak in early 2023 before declining towards the end of the study period. Hospital SARS-CoV-2 detection showed a different pattern, moving from near saturation (100% positivity) in late 2022 to gradually declining positivity throughout the study period. These patterns were consistent with observed patterns in mean viral copies over time by setting (Supplemental Figure 2). We also evaluated neighborhood-level SARS-CoV-2 signal (positivity and viral load) over time (Supplemental Figure 3), which demonstrated a high-degree of intra-neighborhood variability.

**Fig 1.**
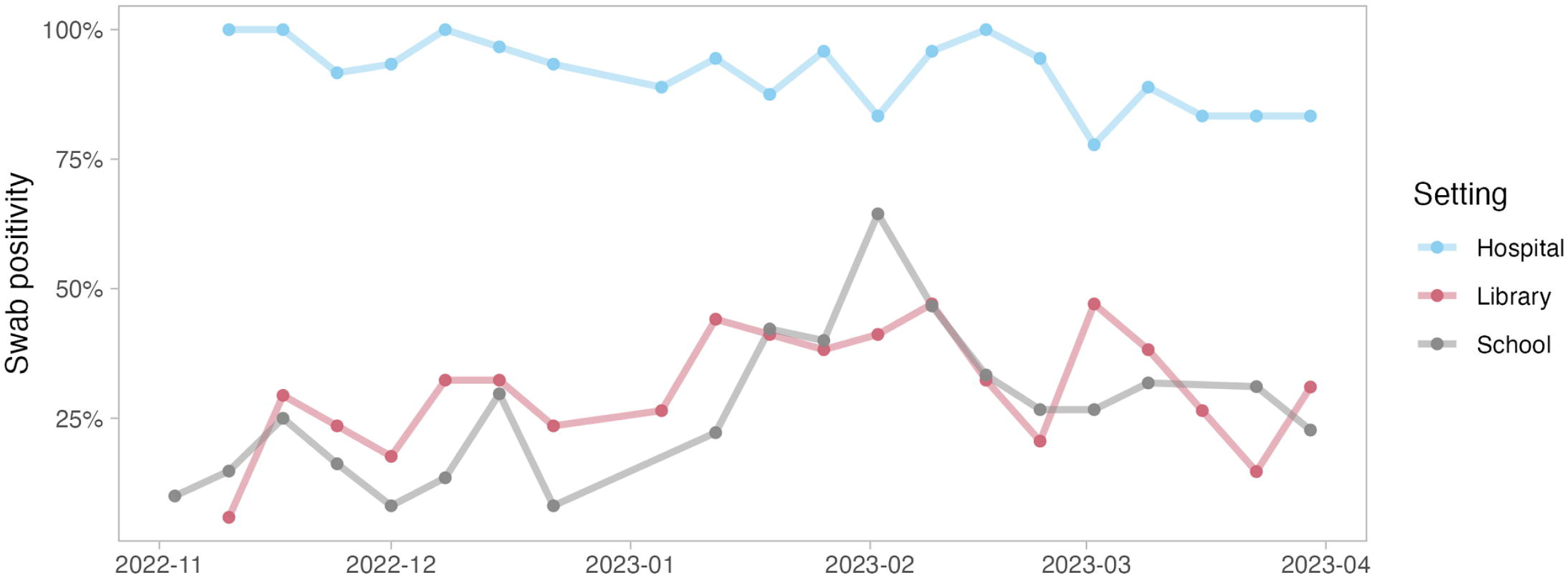
Swab positivity in hospitals, libraries, and schools over the study period.

### Correlation Between SARS-CoV-2 Detection and Outcomes

In order to assess the extent to which overall built environment SARS-CoV-2 detection across all settings predicted relevant burden outcomes, we correlated aggregate built environment SARS-CoV-2 detection with city-level wastewater signal and with hospital COVID-19 burden (admitted COVID-19 patient census). We found no correlation between built environment positivity and wastewater (Spearman’s *r* = 0.15*, p =* 0.5) or between built environment positivity and hospital burden (Spearman’s *r* = 0.13, *p* = 0.6; Figure 2); however, the number of viral copies detected from the built environment was strongly positively correlated with regional hospital burden (Spearman’s r = 0.64, p = 0.0017; Supplemental Table 2), but showed no correlation to regional wastewater (Spearman’s r = - 0.004, p = 0.99). Notably, we also found no correlation between wastewater and hospital burden (Spearman’s *r* = -0.15, *p* = 0.5).

**Fig 2.**
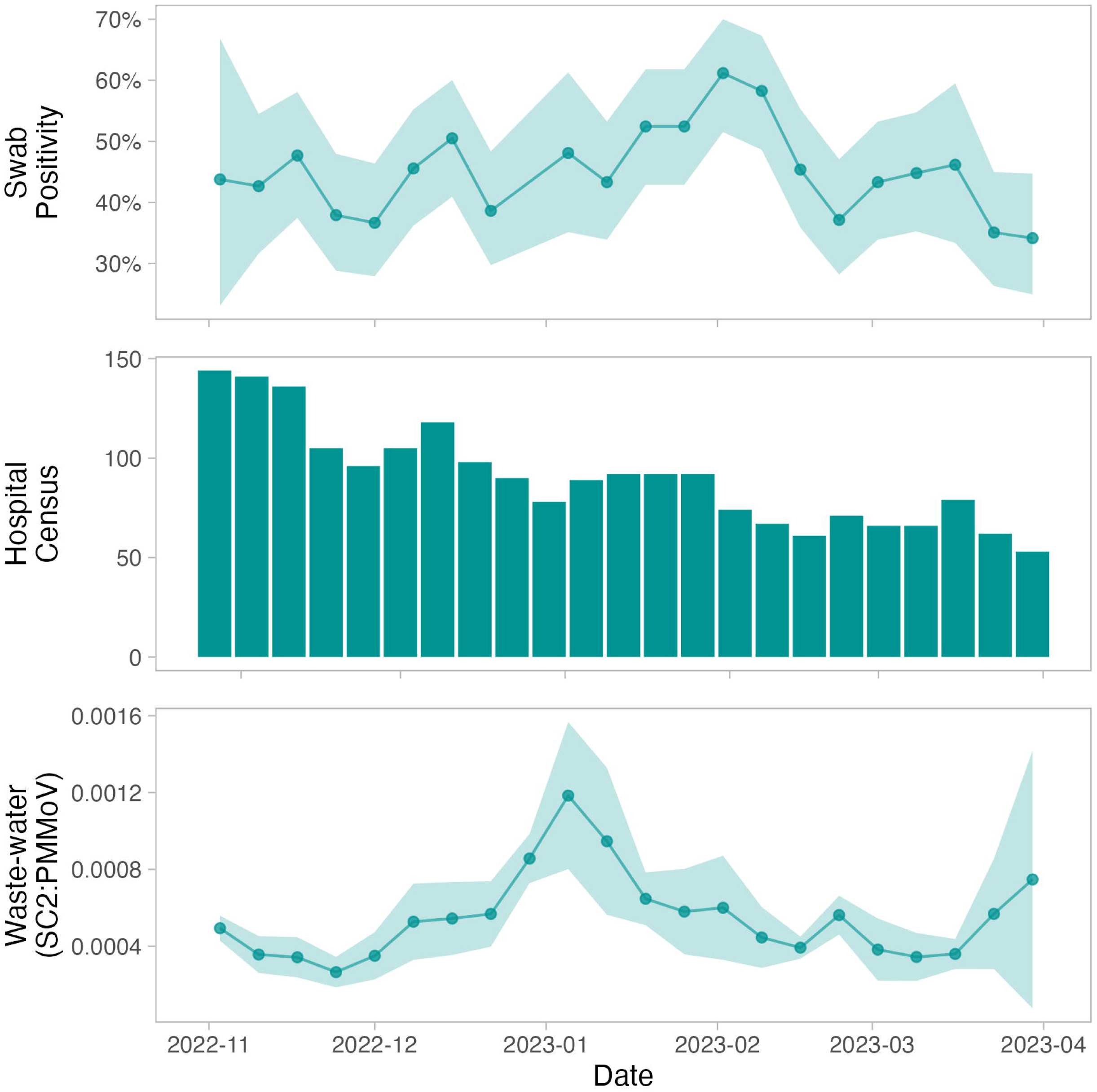
Aggregate environmental swab positivity alongside hospital COVID-19 admitted census across five hospitals included in this study, and weekly regional wastewater signal over the study period. Shaded ribbons show the 95% CI on the weekly means. SC2 = SARS-CoV-2, PMMoV = Pepper mild mottle virus.

We also evaluated the correlation between individual locations/populations and their most relevant burden measure. For hospitals, given the high positivity, we evaluated the relationship between hospital floor SARS-CoV-2 viral load (which is less impacted by loss of trend when 100% of samples are positive) and hospital burden, and found a strong and significant correlation (Spearman’s *r* = 0.76, *p* = 9x10^-5^) (Figure 3). For libraries and schools, we evaluated the relationships between built environment viral copies and both regional wastewater viral copies (Figure 4) as well as hospital burden (Supplemental Table 5), and for both found non-significant correlations with Spearman’s *r* = 0.3 (*p* = 0.18) and Spearman’s *r* = -0.41 (*p*=0.067) respectively.

**Fig 3.**
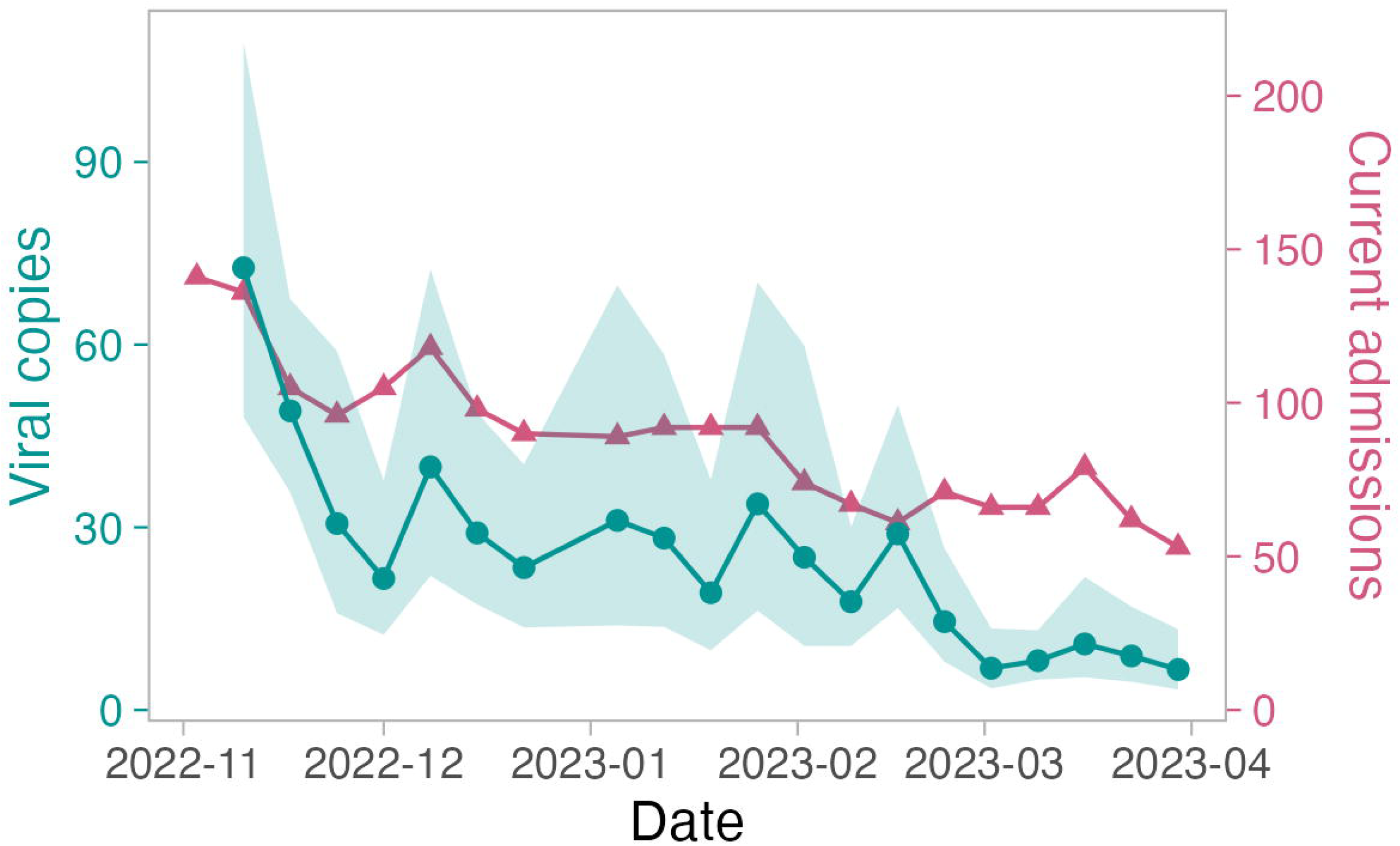
Mean (geometric) of SARS-CoV-2 viral copies (circle) from hospital floor swabs and weekly median number of currently-admitted COVID-19 (census) patients (triangle) across five participating hospitals over the study period. Shading indicates the 95% confidence interval of the mean. ‘Viral copies’ refers to the weekly log_10_-transformed geometric mean number of viral copies plus one copy.

**Fig 4.**
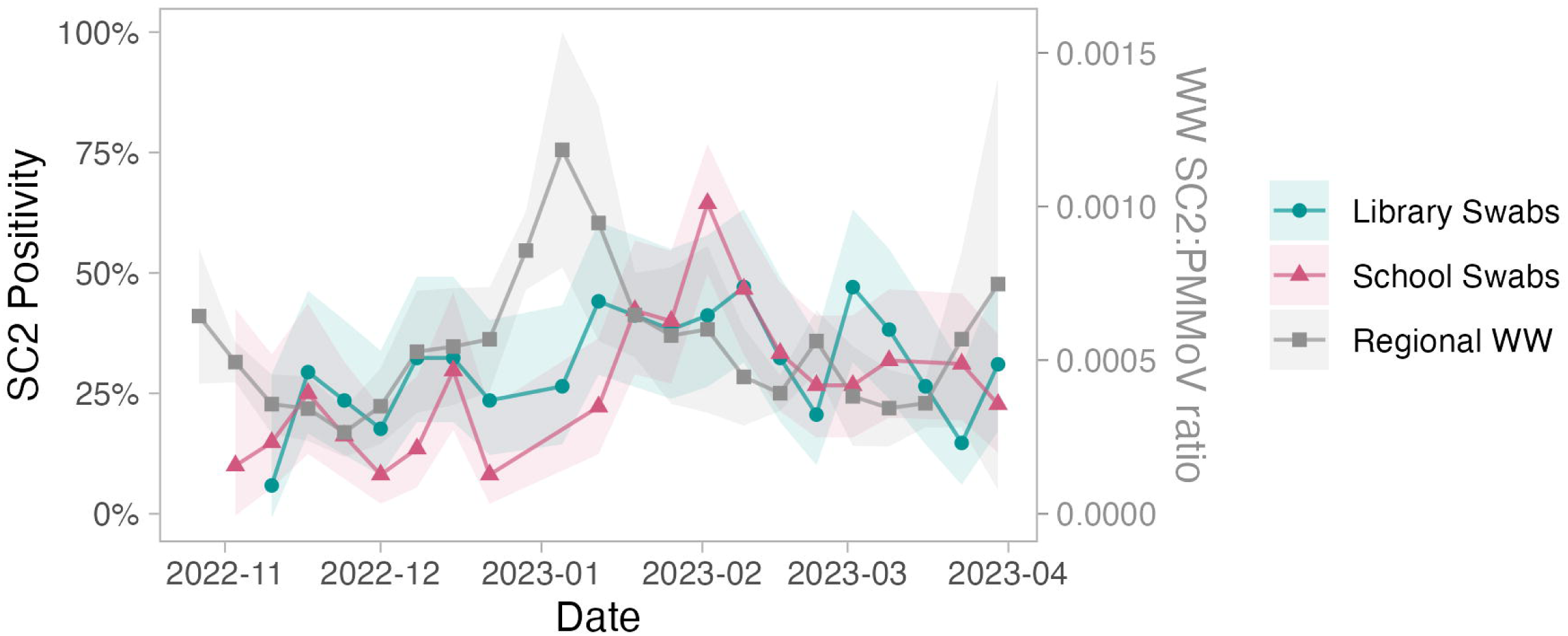
Mean (geometric) of viral copies from libraries and schools compared with regional SARS-CoV-2 wastewater signal over the study period. Shaded areas indicate 95% confidence intervals. ‘Viral copies’ refers to the weekly log_10_-transformed geometric mean number of viral copies plus one copy. WW = wastewater, SC2 = SARS-CoV-2, PMMoV = Pepper mild mottle virus.

A summary of correlations between different environmental swabs and outcomes, including with lagged periods, are shown in Supplemental Table 5. Notably, positive correlations between built environment detection and hospital burden were driven by strong positive correlations with hospital detection but not school or library detection.

Lastly, we evaluated the predictive ability of individual hospital level SARS-CoV-2 measurements (positivity and viral copies) as predictors of hospital burden using a longitudinal fixed-effects linear model. Each model specification was evaluated through cross validation and performance estimated in terms of root-mean-squared error (RMSE) and the coefficient of determination (R^2^) (Supplemental Table 3). For the model using swab test positivity as the predictor, the estimated RMSE was 13.7 ± 0.2 patients and the estimated R^2^ was 0.133 ± 0.011 (mean ± SEM). When swab viral load was used as the predictor, the estimated RMSE decreased to 12.6 ± 3 patients (mean ± SEM) and the estimated R^2^ increased to 0.417 ± 0.017. Including both predictors did not further improve the model performance beyond using viral load alone (RMSE = 12.4 ± 0.27; R^2^ = 0.403 ± 0.021). We fit the final random intercepts model by restricted maximum likelihood (REML) with weekly average viral load as the sole fixed effect. The final model, estimated from 77 weekly observations across five hospitals (combined), had a main effect coefficient of 4.1 (95% CI: 0.15-8.1) for viral load (increase in weekly census 4 for each order of magnitude increase in VL), which indicated a strong and significant association between hospital environmental swab viral loads and hospital burden over time. The standard deviation for sites’ random intercepts was 13. The residuals had a standard deviation of 5.9. Coefficients and standard errors of the three model structures (Swab positive, viral load, and swab positivity + viral load) are shown in Supplemental Table 4.

## DISCUSSION

Built environment testing has recently been evaluated as a spatially resolved method of environmental surveillance with localized clinical and public health implications. Whether built environment surveillance could serve as an adjunct to, or in some instances a replacement for, either traditional surveillance or wastewater surveillance across larger populations and regions has not yet been evaluated. In this study, we performed a prospective city-wide surveillance of built environment detection of SARS-CoV-2, across multiple settings and populations, and correlated these with measures of COVID-19 burden. We found that hospital built environment SARS-CoV-2 positivity and viral load were strongly correlated with COVID-19 hospital census in the region, whereas regional wastewater was not. These findings position built environment surveillance as a potential method to quantify infectious burden amongst institutionalized groups—a burden which may be less well-captured by regional population-wide wastewater surveillance in the context of mixed population immunity.

This study represents a novel evaluation of comprehensive built environment detection for regional surveillance. Previous studies have looked at individual healthcare institutions, university campuses, and schools, though they were inherently limited in the breadth of sampling.^5–7,9^ Our findings that built environment screening provides additional information on regional institutional burden align with prior work conducted in a single health system, which demonstrated that built environment SARS-CoV-2 detection parallels COVID-19 cases.^9^ Notably, in a sensitivity analysis of lagged relationships between built environment detection and hospital burden, and between wastewater and hospital burden, we did not find evidence that lagging improved correlations.

Interestingly, we found little correlation between school and library positivity and regional wastewater signal. This is in keeping with recent work that suggests built environment positivity in schools does not parallel regional wastewater positivity or pediatric COVID-19 hospitalizations.^8^ It is plausible that elementary-aged children and public library patrons do not reflect the major drivers of wastewater signal. This possibility is supported in part by a meta-analysis which suggests there may be differences in SARS-CoV-2 shedding in feces in children compared to adults, including a shorter duration of viral shedding in children.^15^ Thus, these findings do not necessarily mean that built environment surveillance in schools or public libraries are inaccurate measures of population specific burden but, rather, might reflect the challenge of accurately measuring the true burden of COVID-19 in these specific populations.

Our study suggests that built environment testing could be useful to generate a summary of COVID-19 burden within an institutionalized subpopulation across a region. We demonstrate that hospital built environment detection accurately detects a decline in COVID-19 burden within hospitals, which was the opposite trend occurring in regional wastewater. This is a clear divergence of relationships seen earlier on in the pandemic, in which wastewater mirrored hospital burden.^16^ While we applied our approach to hospitals, it may also apply to other settings such as nursing homes and congregate living. Regional built environment viral detection could be helpful both for the current pandemic and future pandemics, to rapidly evaluate burden within important population subgroups that are experiencing less comprehensive or systematic individual testing over time. Moreover, this approach has the potential to provide higher resolution surveillance within facilities to help guide localized infection prevention and control approaches. With increasing evidence that other viruses are detectable through built environment surface surveillance,^17^ this approach may prove useful across a variety of pathogens.

Our study has some limitations. First, we are limited in our ability to collect the most meaningful outcomes for both school-aged children and populations attending public libraries. This is linked to the decline in SARS-CoV-2 testing rates, with human cases at the time predominantly detected via self-performed rapid antigen testing and often not centrally collected through health datasets. We have attempted to correlate our detection with a broader measurement of COVID-19 burden in the general population, namely wastewater signal, but as noted this outcome is not a subgroup-specific measure.

Second, we are limited by our observational design to tease apart whether measurement approaches provide complementary information. For example, the swab signal (positivity and viral load) were similar between school and library populations, which may be a function of similar dynamics in these populations. Using our approach, we are unable to determine which is the case unless we have both a reliable population level metric and the population effects are divergent. Third, we are limited to using normalized wastewater SARS-CoV-2 indices, which prevents our evaluation of relationships with absolute quantitation. Lastly, our evaluation was limited to a six-month period in a given region (Ottawa) and during a time when hospital cases were generally declining, and thus these specifications should be taken into consideration when attempting to generalize these results to longer term or more variable relationships between SARS-CoV-2 levels/detection and regional burden measures, or for other regions.

In conclusion, built environment detection could provide a measure of COVID-19 burden for regional populations of interest, including institutionalized high-risk individuals. Further work to evaluate the use of built environment surveillance for new pathogens, and for at-risk communities without wastewater infrastructure, are needed.

## CONTRIBUTORS

All authors contributed to the conceptualization and design of the project. All authors contributed to the execution of the project, including sample site planning, collection, and laboratory analyses. DRM, JM, RK, and MF contributed to the data analysis, and have accessed/verified the data. DRM and JM drafted the manuscript. All authors contributed to the critical review and revision of the manuscript.

All authors have read and approved the manuscript.

## DATA SHARING STATEMENT

Results of environmental surveillance by swabbing location, and hospital specific COVID-19 census data, cannot be shared due to data and privacy agreements. Ottawa regional waste-water surveillance data has been collected and published by Delatolla *et al.* and is available at their Zenodo repository: https://zenodo.org/records/10794558.

## DECLARATIONS

MF was a consultant for ProofDx, a start up company creating a point of care diagnostic test for COVID-19 and is advisor for SIGNAL1, a start-up company deploying machine learned models to improve inpatient care. ED works for DNA Genotek that provided sampling swabs in-kind for this study in an unrestricted fashion. DNA Genotek had no control over the findings, interpretations, or conclusions published in this paper. All other authors have no conflicts of interest to declare.

## Data Availability

Results of environmental surveillance by swabbing location, and hospital specific COVID-19 census data, cannot be shared due to data and privacy agreements. Ottawa regional waste-water surveillance data has been collected and published by Delatolla et al. and is available at their Zenodo repository: https://zenodo.org/records/10794558.

## ACKNOWLEDGEMENTS

We would like to thank members from the Ottawa Public Library (OPL) and our school board partners for their support in completing this project.

