## Supplementary Materials for "City-wide Built Environment SARS-CoV-2 Detection for COVID-19 Surveillance"

### SUPPLEMENTAL TABLES

- **Supplemental Table 1**. Descriptive statistics by neighborhood.
- **Supplemental Table 2.** Correlation coefficients between geometric means of viral copies
- and hospital COVID-19 admitted patient census or regional wastewater signal, by setting.
- **Supplemental Table 3.** Performance estimates of random intercepts linear models for weekly COVID-19-linked hospital census, evaluated by 5-fold cross-validation repeated 10 times, with predictive performance expressed in terms of root mean squared error (RMSE) and $R^2$.
- **Supplemental Table 4.** Coefficients, standard errors, and other information pertaining to finalized mixed effects linear models for weekly hospital census with each or both surface-detection variables as predictors (models which were evaluated by cross-validation in Supplemental Table 3).
- **Supplemental Table 5.** Spearman and Pearson correlation tests from surface detection, waste-water surveillance and hospital census variables.

### SUPPLEMENTAL FIGURES

- **Supplemental Figure 1.** Neighborhoods evaluated in this study.
- **Supplemental Figure 2.** Geometric mean of viral copies over time by population.
- **Supplemental Figure 3.** Neighborhood-level aggregated SARS-CoV-2 positivity and viral copies over time.

**SUPPLEMENTAL TABLES**

**Supplemental Table 1**. Descriptive statistics by neighborhood. 'Viral copies' refers to the weekly $log_{10}$-transformed geometric mean number of viral copies plus one copy.

| Area | Swabs (n) | Positivity (95% CI) | Viral Copies (95% CI) | Start Date | End Date | Sites (N) |
|---|---|---|---|---|---|---|
| Neighborhood-1 | 418 | 46% (41-51) | 2.82 (2.49-3.19) | 2022-11-03 | 2023-03-30 | 3 |
| Neighborhood-2 | 100 | 20% (13-29) | 1.43 (1.23-1.66) | 2022-11-09 | 2023-03-30 | 1 |
| Neighborhood-3 | 336 | 50% (45-55) | 4.72 (3.84-5.8) | 2022-11-09 | 2023-03-30 | 3 |
| Neighborhood-4 | 348 | 54% (48-59) | 4.96 (4.11-5.99) | 2022-11-11 | 2023-03-30 | 3 |
| Neighborhood-5 | 424 | 41% (36-46) | 2.88 (2.49-3.33) | 2022-10-25 | 2023-03-30 | 3 |
| Neighborhood-6 | 237 | 45% (39-51) | 2.97 (2.47-3.58) | 2022-11-09 | 2023-03-30 | 3 |

**Supplemental Table 2.** Correlation coefficients between geometric means of viral copies and hospital COVID-19 admitted patient census or regional wastewater signal, by setting. Viral copies refers to the weekly $log_{10}$-transformed geometric mean number of viral copies plus one copy.

| **Variables** | **Spearman r (p-value)** | **Pearson r (p-value)** |
|---|---|---|
| Swab viral copies ~ hospital census | 0.643 ($1.7 \times 10^{-3}$) | 0.728 ($1.9 \times 10^{-4}$) |
| Swab viral copies ~ regional waste-water | -0.004 (0.99) | 0.074 (0.8) |

**Supplemental Table 3.** Performance estimates of random intercepts linear models for weekly COVID-19-linked hospital census, evaluated by 5-fold cross-validation repeated 10 times, with predictive performance expressed in terms of root mean squared error (RMSE) and $R^2$. Viral copies represent the weekly $\log_{10}$-transformed geometric mean number of viral copies plus one copy.

| **Fixed Effect Specification** | **RMSE (patients)** | **$R^2$** |
|---|---|---|
| Positivity | 13.73 ± 0.25 | 0.133 ± 0.011 |
| Viral copies | 12.57 ± 0.26 | 0.417 ± 0.017 |
| Positivity + Viral copies | 12.41 ± 0.27 | 0.403 ± 0.021 |

**Supplemental Table 4.** Coefficients, standard errors, and other information pertaining to finalized mixed effects linear models for weekly hospital census with each or both surface-detection variables as predictors (models which were evaluated by cross-validation in Supplemental Table 3). All models included the same number of observations (N=77 weekly observations) from 5 groups (ie. a random intercept for each hospital). Uncertainty intervals (equal-tailed) and p-values (two-tailed) computed using a Wald t-distribution approximation. Uncertainty intervals for random effect variances computed using a Wald z-distribution approximation. Weekly viral copies values were computed as the $log_{10}$-transformed geometric mean of the number of viral copies plus one copy (to avoid undefined values for negative tests).

| Model Predictors | Fixed or Random Effect | Parameter | Coef | SE | 95% CI | t (df) | p |
|---|---|---|---|---|---|---|---|
| Viral copies | Fixed | Intercept | 13.2 | 6.72 | -0.2, 26.59 | 1.96 (73) | 0.053 |
| | Fixed | Viral copies | 4.12 | 2.03 | 0.09, 8.16 | 2.04 (73) | 0.045 |
| | Random | SD (Intercept: site) | 13.45 | 4.85 | 6.64, 27.26 | - | - |
| | Random | SD (Residual) | 5.95 | 0.50 | 5.05, 7.01 | - | - |
| Positivity | Fixed | Intercept | 19.93 | 8.68 | 2.63, 37.22 | 2.30 (73) | 0.025 |
| | Fixed | Positivity | -0.87 | 5.95 | -12.73,10.99 | -0.15 (73) | 0.88 |
| | Random | SD (Intercept: site) | 14.76 | 5.29 | 7.31, 29.82 | - | - |
| | Random | SD (Residual) | 6.09 | 0.51 | 5.16, 7.17 | - | - |
| Positivity + viral copies | Fixed | Intercept | 21.27 | 8.1 | 5.13, 37.42 | 2.63 (72) | 0.011 |
| | Fixed | Viral copies | 6.82 | 2.52 | 1.8, 11.84 | 2.71 | 0.008 |
| | Fixed | Positivity | -12.73 | 7.23 | -27.14, 1.68 | -1.76 | 0.082 |
| | Random | SD (Intercept: site) | 13.4 | 4.83 | 6.61, 27.15 | - | - |
| | Random | SD (Residual) | 5.86 | 0.5 | 4.97, 6.92 | - | - |

**Supplemental Table 5.** Spearman and Pearson correlation tests from surface detection, waste-water surveillance and hospital census variables. Correlations between environmental detection variables with lagging outcomes were also tested where noted (e.g.: *Admits n weeks later*). Subgroups of environmental detection measures are indicated where values were taken from hospitals, schools, libraries, or schools and libraries, as opposed to from all sites. Variable names have been abbreviated as follows: "Census" represent the total number of COVID-19-related hospital census across the five hospitals monitored by surface sampling; "Copies" represents the weekly $\log_{10}$-transformed geometric mean number of viral copies plus one copy; "WW" represents the weekly mean value of SARS-CoV-2 N1 and N2 targets, relative to PPMoV.

| Environmental detection | Outcome | Spearman | | Pearson | |
|---|---|---|---|---|---|
| | | r | p-value | r | p-value |
| Copies: Hospital | Census | 0.76 | 9.5e-05 | 0.84 | 3.9e-06 |
| Copies: Hospital | Census 1 week later | 0.65 | 0.0025 | 0.59 | 0.0073 |
| Copies: Hospital | Census 2 weeks later | 0.56 | 0.016 | 0.53 | 0.022 |
| Copies: Library | Census | -0.14 | 0.57 | -0.3 | 0.21 |
| Copies: Library | Census 1 week later | -0.16 | 0.51 | -0.28 | 0.25 |
| Copies: Library | Census 2 weeks later | -0.18 | 0.47 | -0.27 | 0.29 |
| Copies: School | Census | -0.58 | 0.0092 | -0.37 | 0.12 |
| Copies: School | Census 1 week later | -0.66 | 0.003 | -0.45 | 0.061 |
| Copies: School | Census 2 weeks later | -0.77 | 0.00028 | -0.61 | 0.0097 |
| Copies: All sites | Census | 0.64 | 0.0017 | 0.73 | 0.00019 |
| Copies: All sites | Census 1 week later | 0.49 | 0.03 | 0.55 | 0.012 |
| Copies: All sites | Census 2 weeks later | 0.34 | 0.16 | 0.31 | 0.19 |
| Copies: Schools & Libraries | Census | -0.41 | 0.065 | -0.44 | 0.046 |
| Copies: Schools & Libraries | Census 1 week later | -0.49 | 0.028 | -0.48 | 0.033 |
| Copies: Schools & Libraries | Census 2 weeks later | -0.58 | 0.0091 | -0.53 | 0.019 |
| Positivity: Hospital | Census | 0.54 | 0.013 | 0.59 | 0.0066 |
| Positivity: Hospital | Census 1 week later | 0.4 | 0.087 | 0.47 | 0.044 |
| Positivity: Hospital | Census 2 weeks later | 0.31 | 0.21 | 0.35 | 0.16 |
| Positivity: Library | Census | -0.25 | 0.3 | -0.4 | 0.084 |
| Positivity: Library | Census 1 week later | -0.27 | 0.27 | -0.3 | 0.21 |
| Positivity: Library | Census 2 weeks later | -0.37 | 0.13 | -0.4 | 0.099 |
| Positivity: School | Census | -0.53 | 0.019 | -0.52 | 0.022 |
| Positivity: School | Census 1 week later | -0.66 | 0.0028 | -0.63 | 0.0047 |
| Positivity: School | Census 2 weeks later | -0.76 | 0.00035 | -0.74 | 0.00076 |
| Positivity: All sites | Census | 0.13 | 0.59 | 0.021 | 0.93 |
| Positivity: All sites | Census 1 week later | -0.16 | 0.5 | -0.21 | 0.38 |
| Positivity: All sites | Census 2 weeks later | -0.43 | 0.067 | -0.46 | 0.045 |
| Positivity: Schools & Libraries | Census | -0.47 | 0.03 | -0.56 | 0.0079 |
| Positivity: Schools & Libraries | Census 1 week later | -0.55 | 0.012 | -0.6 | 0.0051 |
| Positivity: Schools & Libraries | Census 2 weeks later | -0.63 | 0.0041 | -0.64 | 0.003 |
| WW | Census | -0.15 | 0.51 | -0.075 | 0.73 |
| WW | Census 1 week later | -0.089 | 0.69 | 0.041 | 0.86 |
| WW | Census 2 weeks later | -0.067 | 0.77 | 0.086 | 0.71 |
| WW 1 week earlier | Census 2 weeks later | -0.071 | 0.77 | 0.0075 | 0.97 |
| WW 1 week later | Census | -0.22 | 0.33 | -0.2 | 0.36 |
| WW 1 week later | Census 1 week later | -0.23 | 0.29 | -0.13 | 0.56 |
| WW 1 week later | Census 2 weeks later | -0.21 | 0.37 | -0.012 | 0.96 |
| Copies: Hospital | WW | -0.11 | 0.65 | -0.077 | 0.75 |
| Copies: Hospital | WW 1 week ago | -0.021 | 0.93 | 0.016 | 0.95 |
| Copies: Hospital | WW 1 week later | -0.051 | 0.84 | -0.16 | 0.51 |
| Copies: Library | WW | 0.2 | 0.4 | 0.079 | 0.74 |

| Environmental detection | Outcome | Spearman | | Pearson | |
|---|---|---|---|---|---|
| | | r | p-value | r | p-value |
| Copies: Library | WW 1 week ago | 0.5 | 0.026 | 0.38 | 0.097 |
| Copies: Library | WW 1 week later | -0.065 | 0.79 | -0.2 | 0.41 |
| Copies: School | WW | 0.3 | 0.21 | 0.21 | 0.38 |
| Copies: School | WW 1 week ago | 0.35 | 0.14 | 0.28 | 0.25 |
| Copies: School | WW 1 week later | 0.19 | 0.45 | -0.037 | 0.88 |
| Copies: All sites | WW | -0.0039 | 0.99 | 0.074 | 0.75 |
| Copies: All sites | WW 1 week ago | 0.29 | 0.21 | 0.22 | 0.34 |
| Copies: All sites | WW 1 week later | -0.18 | 0.45 | -0.14 | 0.56 |
| Copies: Schools & Libraries | WW | 0.3 | 0.18 | 0.14 | 0.54 |
| Copies: Schools & Libraries | WW 1 week ago | 0.45 | 0.038 | 0.32 | 0.16 |
| Copies: Schools & Libraries | WW 1 week later | 0.14 | 0.56 | -0.08 | 0.74 |
| Positivity: Hospital | WW | -0.26 | 0.26 | -0.15 | 0.53 |
| Positivity: Hospital | WW 1 week ago | -0.12 | 0.61 | -0.088 | 0.71 |
| Positivity: Hospital | WW 1 week later | -0.21 | 0.39 | -0.13 | 0.61 |
| Positivity: Library | WW | 0.25 | 0.29 | 0.17 | 0.47 |
| Positivity: Library | WW 1 week ago | 0.59 | 0.0057 | 0.49 | 0.027 |
| Positivity: Library | WW 1 week later | -0.021 | 0.93 | -0.12 | 0.61 |
| Positivity: School | WW | 0.26 | 0.29 | 0.18 | 0.47 |
| Positivity: School | WW 1 week ago | 0.37 | 0.12 | 0.24 | 0.32 |
| Positivity: School | WW 1 week later | 0.14 | 0.59 | -0.067 | 0.79 |
| Positivity: All sites | WW | 0.15 | 0.51 | 0.088 | 0.71 |
| Positivity: All sites | WW 1 week ago | 0.41 | 0.066 | 0.32 | 0.16 |
| Positivity: All sites | WW 1 week later | 0.11 | 0.64 | -0.096 | 0.69 |
| Positivity: Schools & Libraries | WW | 0.26 | 0.26 | 0.15 | 0.51 |
| Positivity: Schools & Libraries | WW 1 week ago | 0.45 | 0.04 | 0.32 | 0.15 |
| Positivity: Schools & Libraries | WW 1 week later | 0.11 | 0.64 | -0.056 | 0.81 |

### SUPPLEMENTAL FIGURES

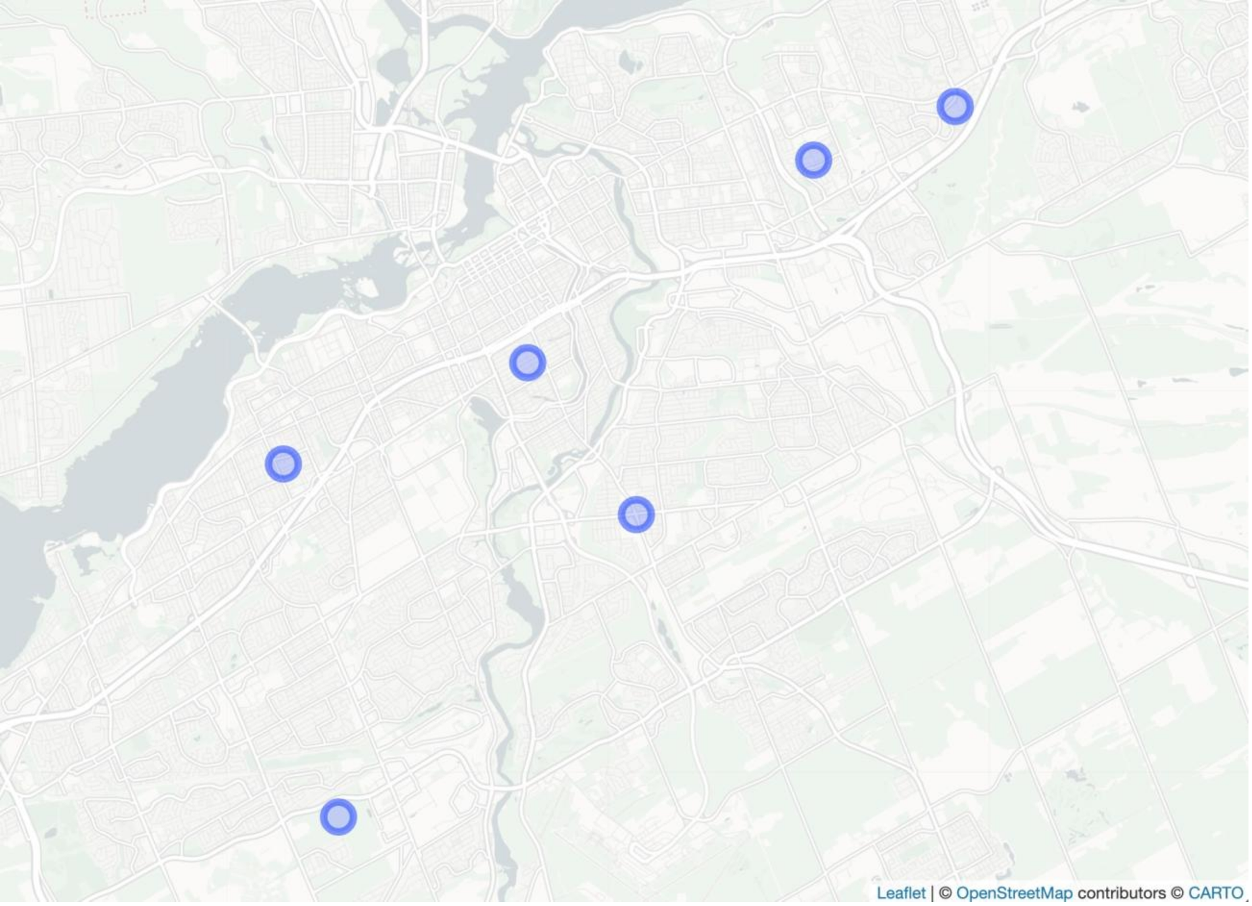


**Supplemental Figure 1.** Neighborhoods evaluated in this study.

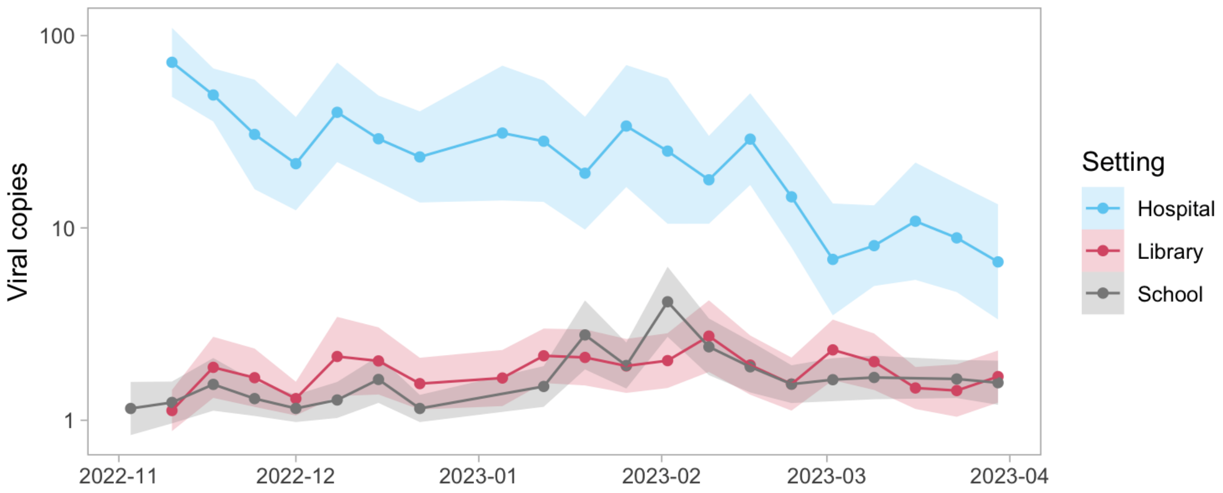


**Supplemental Figure 2.** Geometric mean of viral copies over time by population.

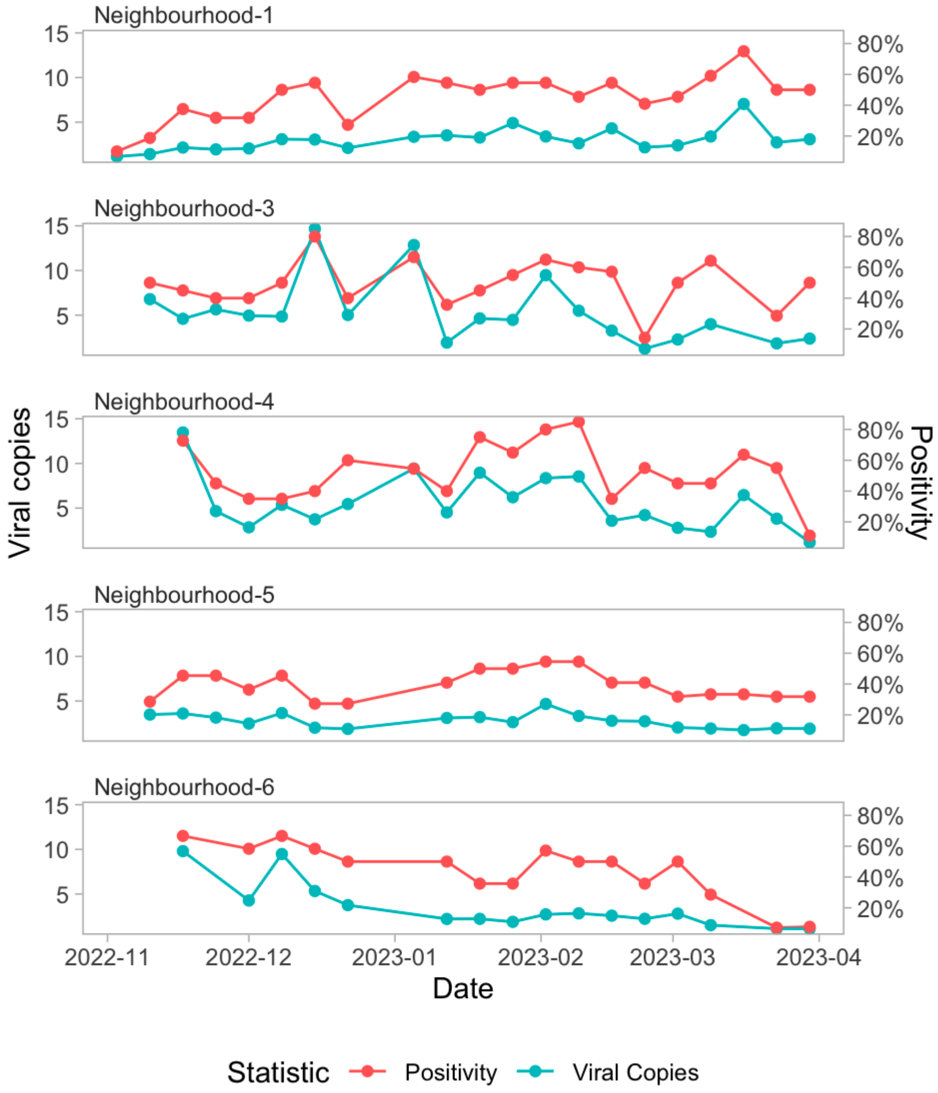


**Supplemental Figure 3.** Neighborhood-level aggregated SARS-CoV-2 positivity and viral copies over time.
